# Approximating vaccine delivery costs to reach zero-dose children: a Bayesian meta-regression analysis

**DOI:** 10.1101/2025.10.05.25337360

**Authors:** Allison Portnoy, Emma Clarke-Deelder, Taylor A. Holroyd, Daniel R. Hogan, Tewodaj Mengistu

**Affiliations:** Department of Global Health, Boston University School of Public Health; Center for Health Decision Science, Harvard T.H. Chan School of Public Health; Department of Epidemiology and Public Health, Swiss Tropical & Public Health Institute; Gavi, the Vaccine Alliance

## Abstract

The Immunization Agenda 2030 calls for reaching all people with immunization services, including ‘zero-dose’ children—children who have not received any routine vaccines. To plan and finance efforts to fully vaccinate these children and improve coverage and equity, decision-makers need reliable cost estimates. However, primary data on the costs of reaching zero-dose children, typically part of disadvantaged and hard-to-reach populations, are scarce. This study approximates these costs using standardized, country-level estimates of vaccine delivery unit costs for outreach delivery in low- and middle-income countries (LMICs). We extracted outreach delivery cost-per-dose estimates for childhood immunization services from the 2024 update of the Immunization Delivery Cost Catalogue. Using these data, we developed a meta-regression model to estimate standardized outreach vaccine delivery unit costs. The generalized linear model assumed a Gamma-distributed outcome with a log link and included both country-level and study-level predictors: study year, economic or financial cost basis, routine or campaign delivery, and full or incremental costing approach. The fitted model was used to estimate 2024 outreach delivery costs per dose for 129 LMICs. The model was estimated using 48 observations from 19 countries focused on outreach or mobile vaccine delivery. The best-fitting specification included diphtheria-tetanus-pertussis (DTP1) coverage, per-capita gross domestic product, and under-five population size as predictors. For 2024, the predicted mean economic cost per dose was $8.65 (95% credible interval $2.33–23.71), averaged across all 129 LMICs. To fully immunize a zero-dose child with 13 recommended vaccinations, the equivalent cost estimate was $112.45 ($30.29–308.23). Reaching zero-dose children is crucial for improving equity in global health, and estimates of the costs of doing so are needed to inform budgeting for immunization programs. These meta-regression-based cost estimates can help countries to improve budgeting, planning, and resource allocation for efforts to reach zero-dose children.

**Key Messages:**

- Despite global commitments under the Immunization Agenda 2030, millions of ‘zero-dose’ children in low- and middle-income countries (LMICs) remain unvaccinated, often due to poverty, geographic inaccessibility, or conflict.
- Given the resource intensity of collecting data on the costs of reaching zero-dose children, existing evidence on is primarily available for a small number of countries from often small-scale implementation studies, providing important context-specific insights while highlighting the need for broader standardized estimates.
- Using a Bayesian meta-regression framework, this study estimated standardized, country-level estimates of vaccine delivery costs based on data for outreach delivery—a proxy for the costs of reaching zero-dose children—resulting in an average 2024 economic cost of $8.65 per dose and $112.45 per fully vaccinated zero-dose child across 129 LMICs.
- This study provides an evidence base to estimate the investments needed to close immunization coverage gaps under the Immunization Agenda 2030.

## Introduction

A significant public health concern in low- and middle-income country (LMIC) settings is the persistence of children who have not received any routine vaccinations—commonly referred to as ‘zero-dose’ children (World Health Organization, 2020). The World Health Organization’s Immunization Agenda 2030 aims to fully vaccinate all people with immunization services, including ‘zero-dose’ children (World Health Organization). Despite decades of progress in improving immunization coverage, the number of children remaining unvaccinated has increased in recent years, leaving them highly susceptible to preventable diseases such as measles, polio, diphtheria, and pertussis (World Health Organization, 2023). This gap in immunization not only contributes to elevated morbidity and mortality rates among unprotected children but also undermines broader efforts to achieve herd immunity and control outbreaks. The presence of zero-dose children is often concentrated in marginalized populations facing systemic barriers such as poverty, conflict, displacement, and weak health infrastructure (Wendt et al., 2022, Hogan and Gupta, 2023).

We have limited direct evidence quantifying the costs of fully vaccinating zero-dose children with needed vaccinations and the evidence that does exist reveals important gaps. For example, Clarke-Deelder et al. (2024) examined India’s Intensified Mission Indradhanush (IMI), and estimated the incremental cost of the campaign at about US$82.99 per zero-dose child reached (with wide uncertainty) from the program perspective, and show that the strategy was likely cost-effective under per-capita GDP thresholds (Clarke-Deelder et al., 2024). Meanwhile, Portnoy et al. (2020) provided ‘standardized’ delivery unit cost estimates across LMICs (excluding vaccine price) showing an average cost of US$1.87 per vaccine dose delivered via routine services, though this does not specifically isolate zero-dose children or special outreach efforts (Portnoy et al., 2020). Additionally, a commentary by Portnoy, et al. (2021) highlights that relatively few immunization cost studies have estimated marginal or scale-up costs or examined how costs vary by coverage levels, reflecting the methodological challenges of measuring the additional investments needed to reach zero-dose populations (Portnoy et al., 2021). More recently, Levin et al. (2024) carried out a scoping review of interventions explicitly aiming to reach zero-dose children in LMICs, identifying eleven such studies; intervention costs ranged widely—from about US$0.08 per additional dose for low-touch strategies like SMS reminders in Kenya to about US$67 per dose for more intensive interventions such as cash transfers in Nicaragua (Levin et al., 2024).

In order to design and budget for targeted interventions to reduce disease burden and advance health equity globally, decision-makers need to know how much it will cost to fully vaccinate zero-dose children with needed vaccinations. The objective of this study was to approximate the costs of reaching zero-dose children by producing standardized country-level estimates of outreach delivery costs for all countries meeting the World Bank’s LMIC classification (World Bank, 2025a). The identified costs were analyzed in a Bayesian meta-regression analysis framework based on the previously published Portnoy et al. (2020) analysis (Portnoy et al., 2020).

## Methods

### Study data

We relied on a publicly available database describing immunization delivery costs in LMIC settings—the Immunization Delivery Cost Catalogue (IDCC) maintained by the Immunization Costing Action Network (ICAN) (Immunization Costing Action Network (ICAN), 2024, Vaughan et al., 2019). The IDCC is an online web catalog and downloadable Excel spreadsheet of immunization delivery cost evidence in LMIC settings, which describes the results of a systematic review of published and grey literature (covering three categories of key words: ‘immunization’ AND ‘cost’ AND ‘delivery’) available between January 2005 and December 2023. From the IDCC, we identified studies that reported delivery costs of outreach and mobile delivery efforts for childhood vaccines to fully vaccinate children up to the age of 15 with needed immunizations. ‘Outreach’ was defined by IDCC as vaccines delivered through outreach or mobile clinics, whether as part of the routine immunization program or through a mass vaccination campaign. Delivery costs included labor, supply chain, capital, and other service delivery costs. ‘Supply chain’ includes costs for cold chain equipment, vehicles, transport, and fuel; ‘other service delivery’ includes costs for program management (i.e., supervision and monitoring), training, social mobilization, and disease surveillance (Brenzel et al., 2015).

For the identified studies, we extracted the study year, estimates of the delivery cost per dose, whether the vaccine was delivered through routine outreach/mobile delivery vs. mass vaccination campaigns [*Routine*], whether the costing was full or incremental [*Full*], and whether the presented costs were from the financial or economic perspective [*Econ*]—i.e., financial costs refer to expenditures or financial outlays whereas economic costs include both financial costs as well as the opportunity cost associated with using inputs in the immunization program as compared to their next best use. *Routine, Full, Econ* are all binary indicator variables with possible values of 0 and 1 indicating the absence (0) or presence (1) of the corresponding characteristic. Where the IDCC did not include a cost per dose (excluding vaccine costs) directly reported by the study, we reviewed the original article and calculated the cost per dose from available data.

The IDCC 2024 update included delivery cost per dose estimates in 2022 US dollars. In order to present estimates in 2024 US dollars, the study values were inflated using local inflation according to the consumer price index and local currency-to-USD exchange rates.

### Model specification

We compiled data on country-level covariates potentially associated (Portnoy et al., 2020) with outreach delivery unit costs: log gross domestic product (GDP) per capita [*log*(*GDP*)], log total under-five population [*log*(*Pop*)], diphtheria-tetanus-pertussis first dose coverage (ranging from 0 (0% coverage) to 1 (100%) coverage) [*DTP*1], log under-five mortality rate (under-five deaths per 1000 live births) [*log*(*U*5*MR*)], log population density (people per square kilometer of land) [*log*(*Density*)], and urban population (percentage of country population living in urban areas) [*Urban*] (World Bank, 2025b, World Health Organization, 2025, United Nations, 2024).

Country-level covariates were compiled for the year of data collection for each costing study and combined with study-level indicators, i.e., *Year, Routine, Full, Econ*. By using a log transformation of specified covariates, we assumed these explanatory factors relate to the outcome on the multiplicative scale rather than linearly (for example, a doubling in per-capita GDP produces a fixed increase in the logged outcome).

We used a Bayesian meta-regression model to regress outreach delivery unit costs against country-level and study-level explanatory variables. Continuous variables were standardized to a mean of zero and standard deviation of one before fitting the regression model. We constructed a prediction model for the intervention cost per dose outcome (*y_i_*), specified as a generalized linear regression model (GLM), assuming a Gamma distributed outcome and a log link function. In addition to study-level predictors, we selected country-level covariates according to the best fit model using minimized Watanabe-Akaike Information Criterion (WAIC). The model specification assumed a Gamma likelihood function for the observed data *y_i_* where the shape parameter *α* described the residual variance:

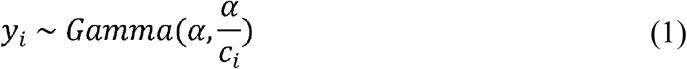

This specification assumed variance proportional to *c_i_*, representing the mean estimate of the delivery cost per dose for a given study. We assumed informative prior distributions for all regression coefficients, which were assumed to follow a normal distribution centered at zero with a standard deviation of 1 (Gelman, 2025). The shape parameter was assumed to follow a half-Cauchy distribution centered at zero with a standard deviation of 5 (Gelman, 2006).

The prediction model was estimated in R software, version 4.4.1, using an adaptive Hamiltonian Monte Carlo algorithm in the Stan software package, version 2.32.6, with four chains of 5000 iterations. The first 2500 iterations were discarded (burn-in period), yielding 10,000 posterior draws for analysis (Hoffman and Gelman, 2014).

Stan model diagnostics (posterior summaries, convergence diagnostics, sampling details, and model fit assessment) were examined to determine any problems encountered by the sampler, and the potential scale reduction factor (i.e., Rhat) for all parameters was evaluated to confirm that the model had successfully converged. We simulated and visually inspected the ability of the model to reproduce the observed distribution of costs.

### Predicted outcomes

The fitted prediction model was used to generate economic outreach delivery cost per dose estimates for each country for the year 2024. As a proxy for the costs of fully vaccinating zero-dose children, we predicted delivery costs assuming routine outreach delivery and a full costing approach. To generate these estimates, we predicted values from the posterior distributions of the fitted model, with covariate values specific to each country and year. We generated aggregated estimates of the cost per dose in all LMICs and in country groups defined by World Bank income level (low-income, lower middle-income, and upper middle-income) by weighting country-specific estimates by the size of the under-five population in each country (World Bank, 2025a). We tested predictive performance by comparing model predictions to the observed cost per dose matched to country and year.

### Sensitivity analysis

We conducted a sensitivity analysis excluding identified outliers to assess the robustness of model estimates to influential observations. Statistical outliers were defined as observations with cost per dose values greater than 1.5 times the interquartile range above the third quartile.

## Results

### Regression model

We identified 22 studies including 48 observations across 19 countries with outreach delivery costs for childhood vaccines reported in the IDCC. Among these studies, the outreach delivery cost per dose ranged from $0.09 to $9.80 in 2024 USD. Table 1 provides summary information on the empirical studies used in the analysis (full details of these studies are provided in Appendix A). In the sample, the average GDP per capita was $1743.57 (standard deviation $1398.95) average population size was 16,700 (std 32,900) and the average diphtheria-tetanus-pertussis (DTP1) coverage was 92.7% (std 6.8%). The best fit model specification was defined as:

**Table 1.** Summary characteristics for outreach delivery unit cost per dose data. Note: LI = low-income; LMI = lower middle-income; SIA = supplementary immunization activities; UMI = upper middle-income.

| Variable | Estimates (*n*) |
| --- | --- |
| Income |  |
| • LI | 24 |
| • LMI | 22 |
| • UMI | 2 |
| Cost type |  |
| • Economic | 27 |
| • Financial | 21 |
| Costing approach |  |
| • Full | 21 |
| • Incremental | 27 |
| Delivery modality |  |
| • Routine | 37 |
| • SIA | 11 |

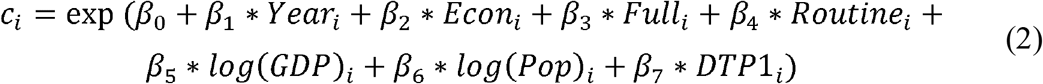

Table 2 reports point estimates and standard errors for regression coefficients and other model parameters. The best fit model demonstrated satisfactory convergence diagnostics, with R-hat values below 1.01 for all estimated parameters and adequate effective sample sizes. Figure 1 displays the in-sample fit comparing observed versus predicted posterior values for the study sample.

**Table 2.** Results for regressions of outreach delivery unit cost per dose on predictors.

| Variable | Mean coefficient (standard error) |
| --- | --- |
| Intercept | -0.158 (0.006) |
| Year | - 0.191 (0.003) |
| Economic cost indicator [*Econ*] | 0.830 (0.003) |
| Full costing indicator [*Full*] | 0.083 (0.005) |
| Routine delivery indicator [*Routine*] | 0.727 (0.004) |
| log(GDP) | 0.126 (0.002) |
| log(Pop) | 0.475 (0.002) |
| DTP1 coverage | 0.525 (0.001) |
| Gamma dispersion parameter alpha | 1.412 |

**Figure 1.**
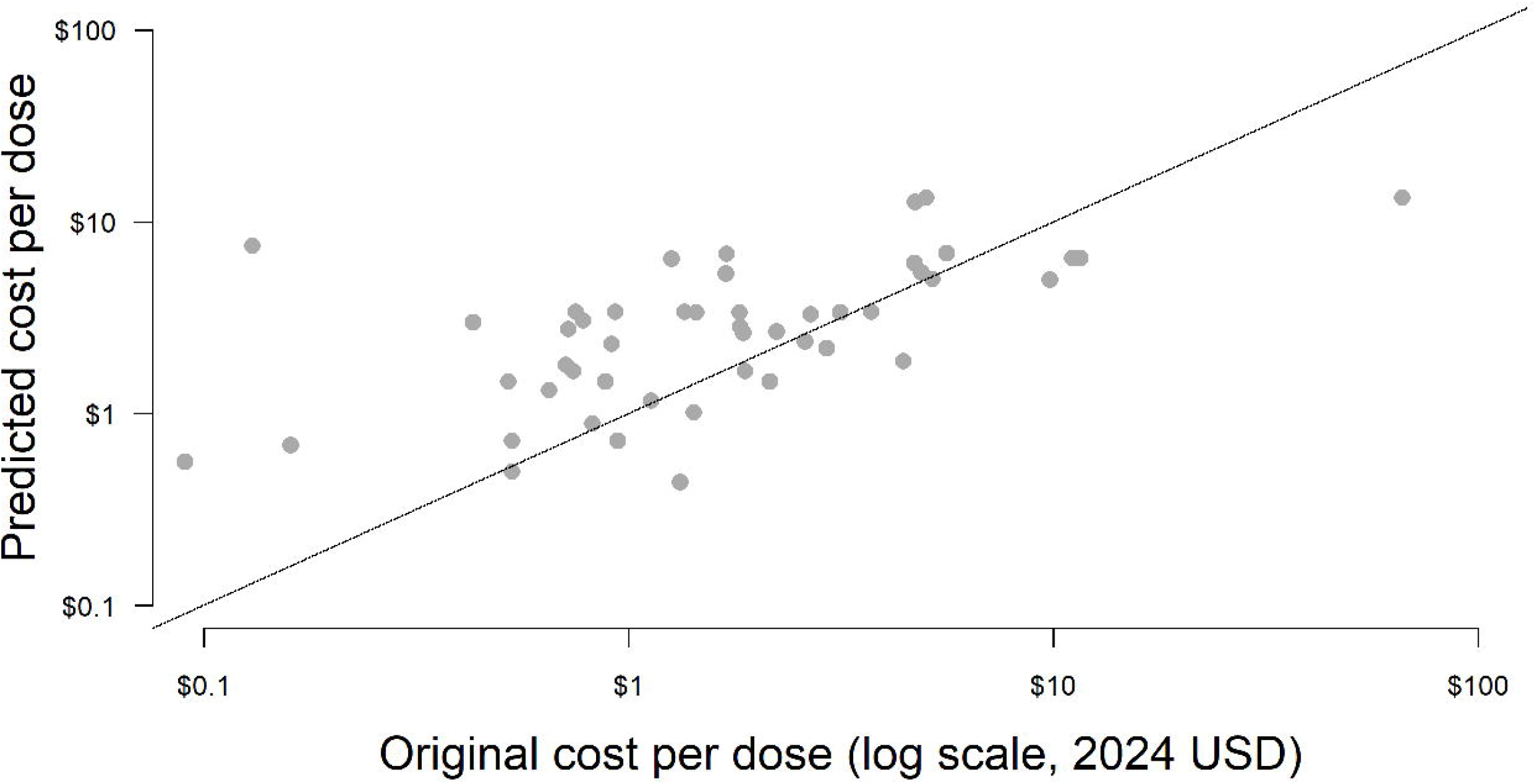
Comparison of predicted cost per dose and published literature cost per dose for outreach delivery.

The fitted model demonstrated increasing costs as price levels (GDP per capita), service delivery volume (under-five population), health system capacity (DTP1 coverage) increased. We calculated first differences to describe the percentage difference in the cost per dose associated with specified changes in individual predictors, holding others fixed. Doubling per-capita GDP would be associated with a 6.7% increase in the cost per dose of outreach-based delivery, whereas doubling the under-five population would be associated with a 34.4% increase. An increase in DTP1 coverage by one percentage point would be associated with a 7.7% increase in the cost per dose of outreach-based delivery. Figure 2 shows how the estimated outreach-based delivery cost per dose varies with coverage, holding all other covariates constant.

**Figure 2.**
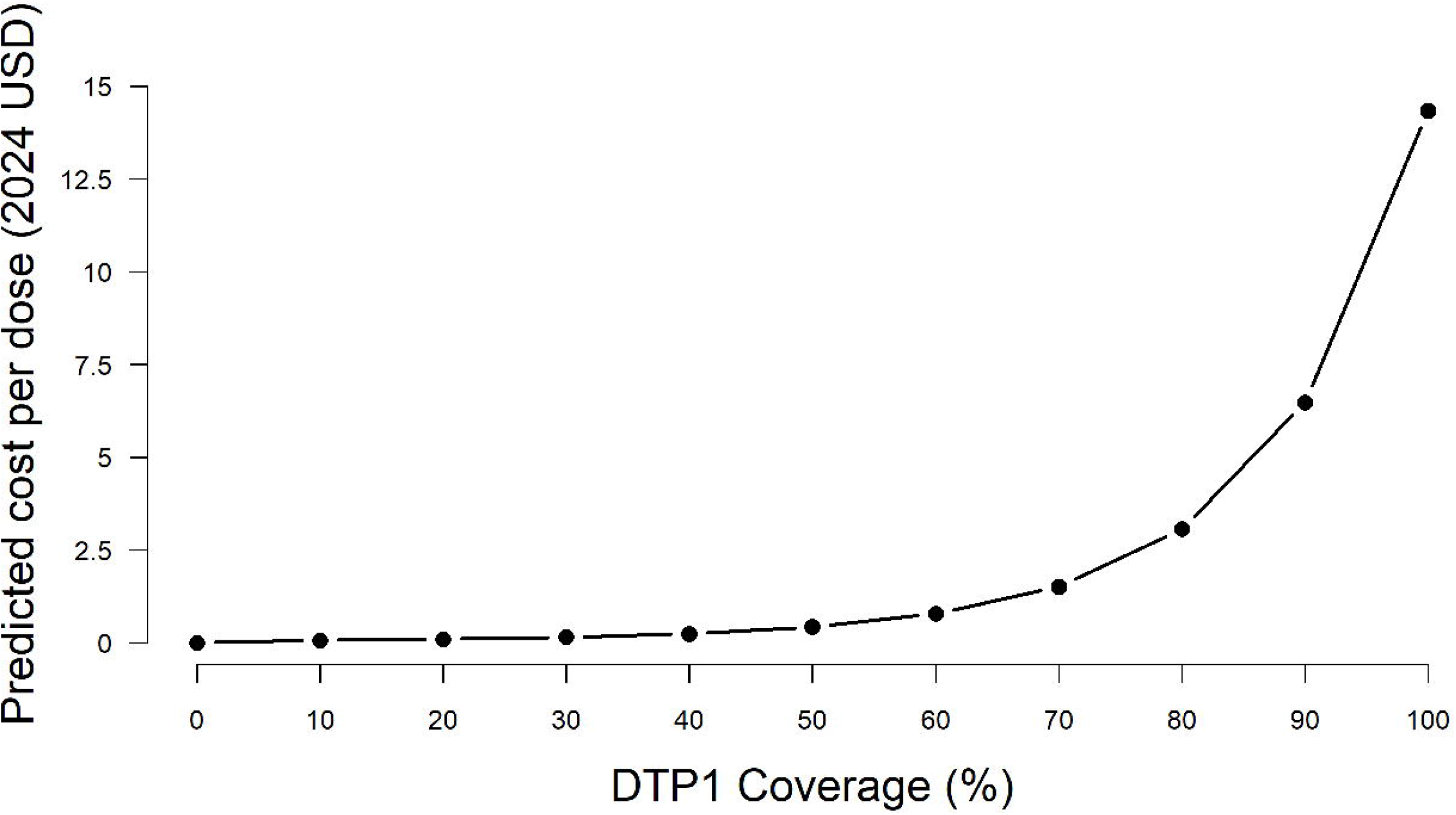
Predicted economic cost per dose in 2024 for routine outreach vaccine delivery by DTP1 (first dose diphtheria-tetanus-pertussis-containing vaccine) coverage for 129 low and middle-income countries.

### Predicted outcomes

For the year 2024, the population-weighted average economic cost per dose was estimated to be $8.65 (95% credible interval $2.33–23.71) for the full costs of routine outreach vaccine delivery across 129 LMICs. By income level, the average predicted economic cost per dose was $2.09 ($0.59–5.47) for low-income countries, $9.91 ($2.59–27.48) for lower middle-income countries, and $10.88 ($2.37–33.93) for upper middle-income countries. Figure 3 presents the country-level cost per dose estimates by GDP per capita and 2025 World Bank income level. The set of country-specific economic cost estimates for outreach delivery cost per dose can be found in Table 3.

**Table 3.**
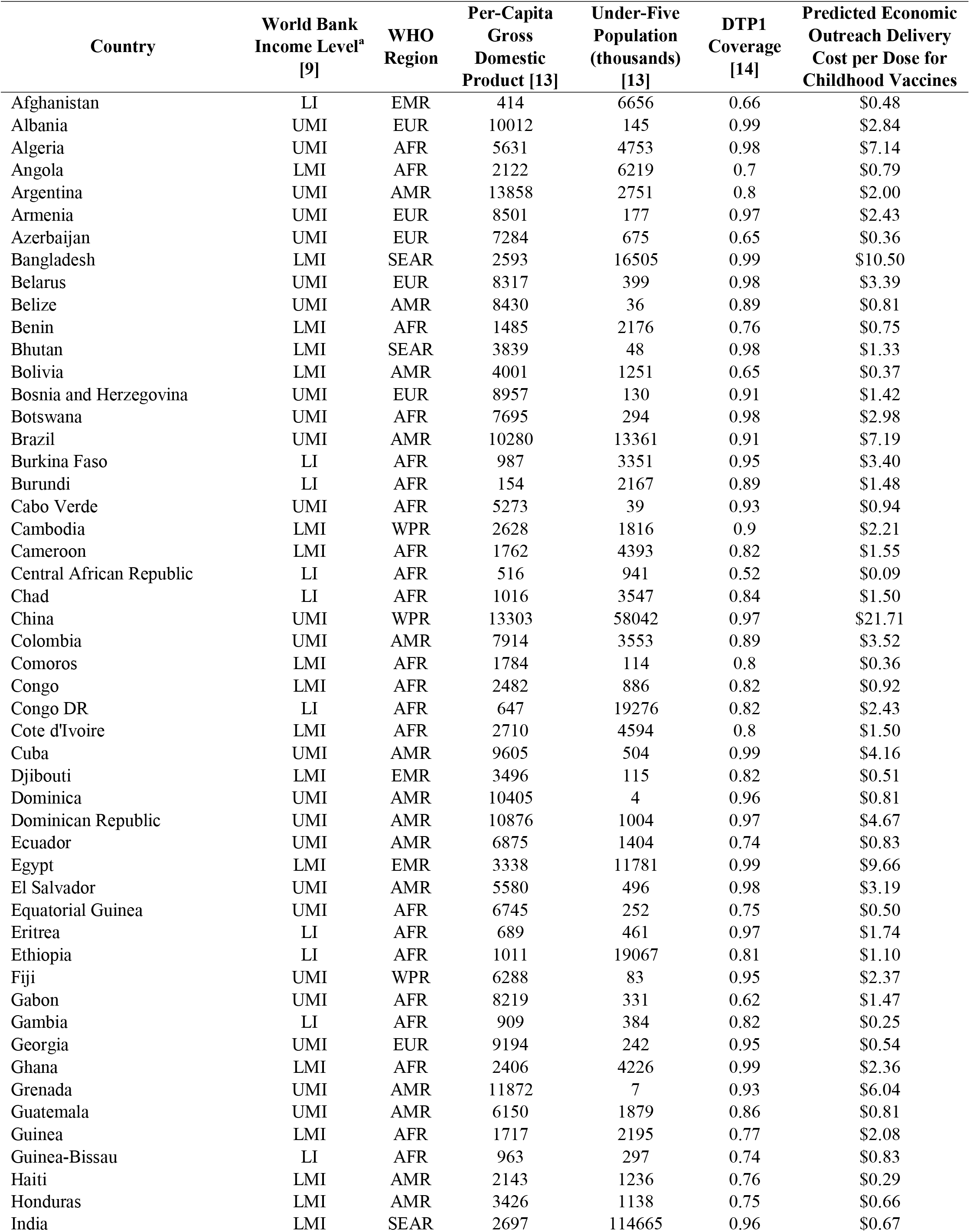
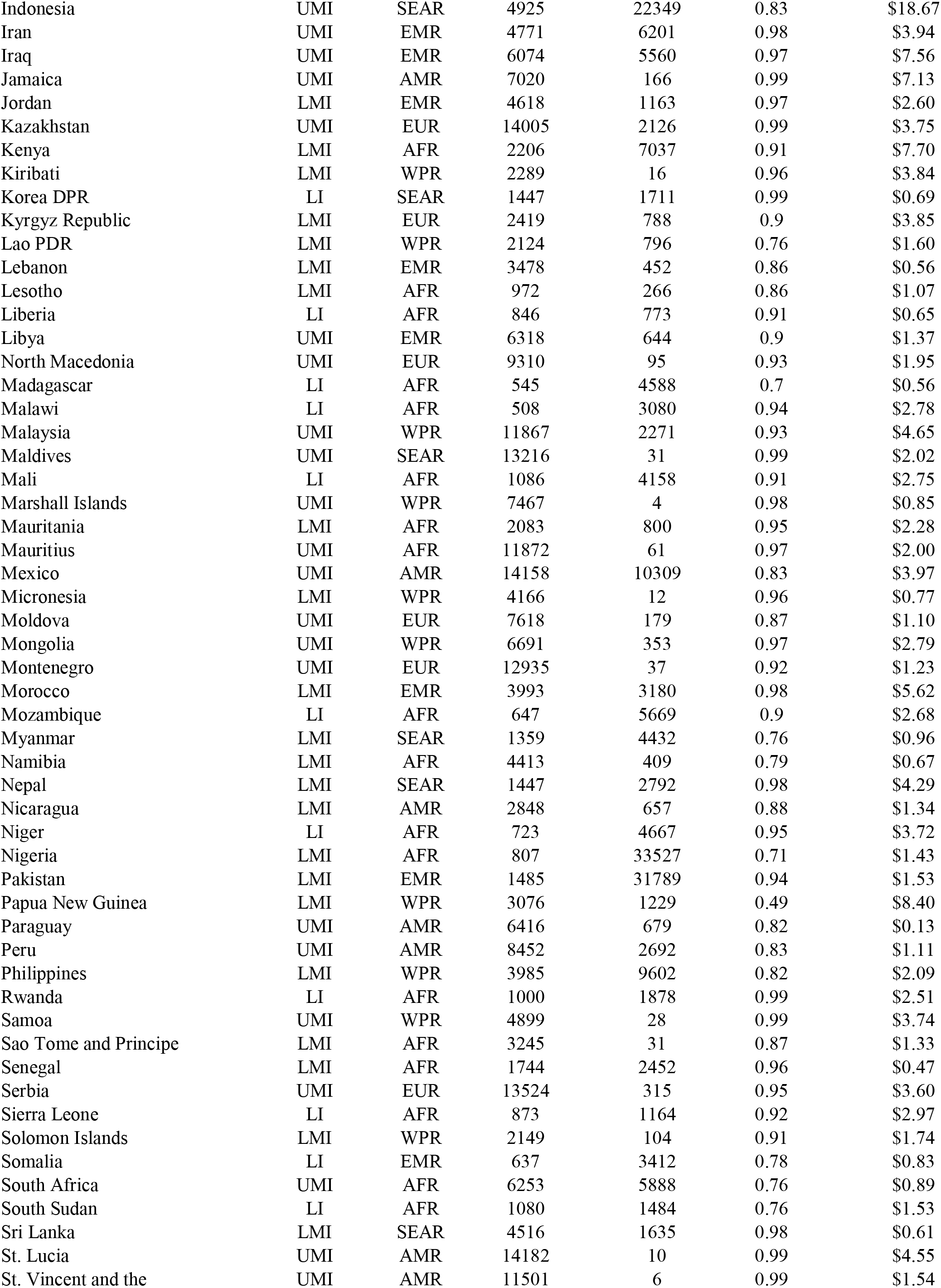
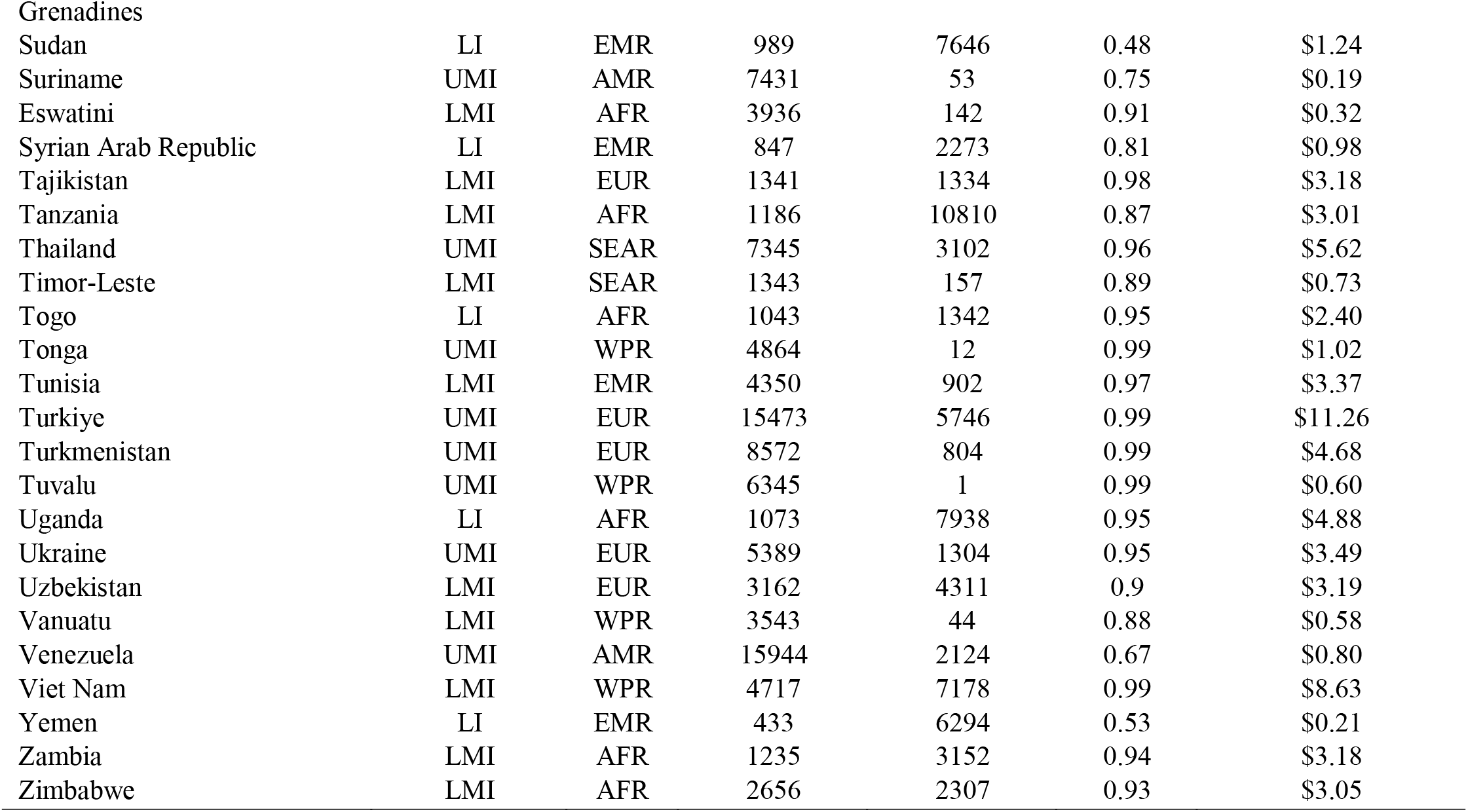
Low- and middle-income country parameters with predicted economic outreach delivery cost per dose in 2024. ^a^ LI: Gross national income (GNI) per capita of $1,135 or less in 2024; LMI: GNI per capita $1,136 and $4,4915; UMI: GNI per capita of $4,496 and $13,935. Note: AFR = African region; AMR = Region of the Americas; DTP1 = diphtheria-tetanus-pertussis first dose coverage; EMR = Eastern Mediterranean region; EUR = European region; LI = low-income; LMI = lower middle-income; SEAR = Southeast Asian region; UMI = upper middle-income; WPR = Western Pacific region.

| Country | World Bank<br>Income Level<sup>a</sup><br>[9] | WHO<br>Region | Per-Capita<br>Gross<br>Domestic<br>Product [13] | Under-Five<br>Population<br>(thousands)<br>[13] | DTP1<br>Coverage<br>[14] | Predicted Economic<br>Outreach Delivery<br>Cost per Dose for<br>Childhood Vaccines |
| --- | --- | --- | --- | --- | --- | --- |
| Afghanistan | LI | EMR | 414 | 6656 | 0.66 | \$0.48 |
| Albania | UMI | EUR | 10012 | 145 | 0.99 | \$2.84 |
| Algeria | UMI | AFR | 5631 | 4753 | 0.98 | \$7.14 |
| Angola | LMI | AFR | 2122 | 6219 | 0.7 | \$0.79 |
| Argentina | UMI | AMR | 13858 | 2751 | 0.8 | \$2.00 |
| Armenia | UMI | EUR | 8501 | 177 | 0.97 | \$2.43 |
| Azerbaijan | UMI | EUR | 7284 | 675 | 0.65 | \$0.36 |
| Bangladesh | LMI | SEAR | 2593 | 16505 | 0.99 | \$10.50 |
| Belarus | UMI | EUR | 8317 | 399 | 0.98 | \$3.39 |
| Belize | UMI | AMR | 8430 | 36 | 0.89 | \$0.81 |
| Benin | LMI | AFR | 1485 | 2176 | 0.76 | \$0.75 |
| Bhutan | LMI | SEAR | 3839 | 48 | 0.98 | \$1.33 |
| Bolivia | LMI | AMR | 4001 | 1251 | 0.65 | \$0.37 |
| Bosnia and Herzegovina | UMI | EUR | 8957 | 130 | 0.91 | \$1.42 |
| Botswana | UMI | AFR | 7695 | 294 | 0.98 | \$2.98 |
| Brazil | UMI | AMR | 10280 | 13361 | 0.91 | \$7.19 |
| Burkina Faso | LI | AFR | 987 | 3351 | 0.95 | \$3.40 |
| Burundi | LI | AFR | 154 | 2167 | 0.89 | \$1.48 |
| Cabo Verde | UMI | AFR | 5273 | 39 | 0.93 | \$0.94 |
| Cambodia | LMI | WPR | 2628 | 1816 | 0.9 | \$2.21 |
| Cameroon | LMI | AFR | 1762 | 4393 | 0.82 | \$1.55 |
| Central African Republic | LI | AFR | 516 | 941 | 0.52 | \$0.09 |
| Chad | LI | AFR | 1016 | 3547 | 0.84 | \$1.50 |
| China | UMI | WPR | 13303 | 58042 | 0.97 | \$21.71 |
| Colombia | UMI | AMR | 7914 | 3553 | 0.89 | \$3.52 |
| Comoros | LMI | AFR | 1784 | 114 | 0.8 | \$0.36 |
| Congo | LMI | AFR | 2482 | 886 | 0.82 | \$0.92 |
| Congo DR | LI | AFR | 647 | 19276 | 0.82 | \$2.43 |
| Cote d'Ivoire | LMI | AFR | 2710 | 4594 | 0.8 | \$1.50 |
| Cuba | UMI | AMR | 9605 | 504 | 0.99 | \$4.16 |
| Djibouti | LMI | EMR | 3496 | 115 | 0.82 | \$0.51 |
| Dominica | UMI | AMR | 10405 | 4 | 0.96 | \$0.81 |
| Dominican Republic | UMI | AMR | 10876 | 1004 | 0.97 | \$4.67 |
| Ecuador | UMI | AMR | 6875 | 1404 | 0.74 | \$0.83 |
| Egypt | LMI | EMR | 3338 | 11781 | 0.99 | \$9.66 |
| El Salvador | UMI | AMR | 5580 | 496 | 0.98 | \$3.19 |
| Equatorial Guinea | UMI | AFR | 6745 | 252 | 0.75 | \$0.50 |
| Eritrea | LI | AFR | 689 | 461 | 0.97 | \$1.74 |
| Ethiopia | LI | AFR | 1011 | 19067 | 0.81 | \$1.10 |
| Fiji | UMI | WPR | 6288 | 83 | 0.95 | \$2.37 |
| Gabon | UMI | AFR | 8219 | 331 | 0.62 | \$1.47 |
| Gambia | LI | AFR | 909 | 384 | 0.82 | \$0.25 |
| Georgia | UMI | EUR | 9194 | 242 | 0.95 | \$0.54 |
| Ghana | LMI | AFR | 2406 | 4226 | 0.99 | \$2.36 |
| Grenada | UMI | AMR | 11872 | 7 | 0.93 | \$6.04 |
| Guatemala | UMI | AMR | 6150 | 1879 | 0.86 | \$0.81 |
| Guinea | LMI | AFR | 1717 | 2195 | 0.77 | \$2.08 |
| Guinea-Bissau | LI | AFR | 963 | 297 | 0.74 | \$0.83 |
| Haiti | LMI | AMR | 2143 | 1236 | 0.76 | \$0.29 |
| Honduras | LMI | AMR | 3426 | 1138 | 0.75 | \$0.66 |
| India | LMI | SEAR | 2697 | 114665 | 0.96 | \$0.67 |
| Indonesia | UMI | SEAR | 4925 | 22349 | 0.83 | \$18.67 |
| Iran | UMI | EMR | 4771 | 6201 | 0.98 | \$3.94 |
| Iraq | UMI | EMR | 6074 | 5560 | 0.97 | \$7.56 |
| Jamaica | UMI | AMR | 7020 | 166 | 0.99 | \$7.13 |
| Jordan | LMI | EMR | 4618 | 1163 | 0.97 | \$2.60 |
| Kazakhstan | UMI | EUR | 14005 | 2126 | 0.99 | \$3.75 |
| Kenya | LMI | AFR | 2206 | 7037 | 0.91 | \$7.70 |
| Kiribati | LMI | WPR | 2289 | 16 | 0.96 | \$3.84 |
| Korea DPR | LI | SEAR | 1447 | 1711 | 0.99 | \$0.69 |
| Kyrgyz Republic | LMI | EUR | 2419 | 788 | 0.9 | \$3.85 |
| Lao PDR | LMI | WPR | 2124 | 796 | 0.76 | \$1.60 |
| Lebanon | LMI | EMR | 3478 | 452 | 0.86 | \$0.56 |
| Lesotho | LMI | AFR | 972 | 266 | 0.86 | \$1.07 |
| Liberia | LI | AFR | 846 | 773 | 0.91 | \$0.65 |
| Libya | UMI | EMR | 6318 | 644 | 0.9 | \$1.37 |
| North Macedonia | UMI | EUR | 9310 | 95 | 0.93 | \$1.95 |
| Madagascar | LI | AFR | 545 | 4588 | 0.7 | \$0.56 |
| Malawi | LI | AFR | 508 | 3080 | 0.94 | \$2.78 |
| Malaysia | UMI | WPR | 11867 | 2271 | 0.93 | \$4.65 |
| Maldives | UMI | SEAR | 13216 | 31 | 0.99 | \$2.02 |
| Mali | LI | AFR | 1086 | 4158 | 0.91 | \$2.75 |
| Marshall Islands | UMI | WPR | 7467 | 4 | 0.98 | \$0.85 |
| Mauritania | LMI | AFR | 2083 | 800 | 0.95 | \$2.28 |
| Mauritius | UMI | AFR | 11872 | 61 | 0.97 | \$2.00 |
| Mexico | UMI | AMR | 14158 | 10309 | 0.83 | \$3.97 |
| Micronesia | LMI | WPR | 4166 | 12 | 0.96 | \$0.77 |
| Moldova | UMI | EUR | 7618 | 179 | 0.87 | \$1.10 |
| Mongolia | UMI | WPR | 6691 | 353 | 0.97 | \$2.79 |
| Montenegro | UMI | EUR | 12935 | 37 | 0.92 | \$1.23 |
| Morocco | LMI | EMR | 3993 | 3180 | 0.98 | \$5.62 |
| Mozambique | LI | AFR | 647 | 5669 | 0.9 | \$2.68 |
| Myanmar | LMI | SEAR | 1359 | 4432 | 0.76 | \$0.96 |
| Namibia | LMI | AFR | 4413 | 409 | 0.79 | \$0.67 |
| Nepal | LMI | SEAR | 1447 | 2792 | 0.98 | \$4.29 |
| Nicaragua | LMI | AMR | 2848 | 657 | 0.88 | \$1.34 |
| Niger | LI | AFR | 723 | 4667 | 0.95 | \$3.72 |
| Nigeria | LMI | AFR | 807 | 33527 | 0.71 | \$1.43 |
| Pakistan | LMI | EMR | 1485 | 31789 | 0.94 | \$1.53 |
| Papua New Guinea | LMI | WPR | 3076 | 1229 | 0.49 | \$8.40 |
| Paraguay | UMI | AMR | 6416 | 679 | 0.82 | \$0.13 |
| Peru | UMI | AMR | 8452 | 2692 | 0.83 | \$1.11 |
| Philippines | LMI | WPR | 3985 | 9602 | 0.82 | \$2.09 |
| Rwanda | LI | AFR | 1000 | 1878 | 0.99 | \$2.51 |
| Samoa | UMI | WPR | 4899 | 28 | 0.99 | \$3.74 |
| Sao Tome and Principe | LMI | AFR | 3245 | 31 | 0.87 | \$1.33 |
| Senegal | LMI | AFR | 1744 | 2452 | 0.96 | \$0.47 |
| Serbia | UMI | EUR | 13524 | 315 | 0.95 | \$3.60 |
| Sierra Leone | LI | AFR | 873 | 1164 | 0.92 | \$2.97 |
| Solomon Islands | LMI | WPR | 2149 | 104 | 0.91 | \$1.74 |
| Somalia | LI | EMR | 637 | 3412 | 0.78 | \$0.83 |
| South Africa | UMI | AFR | 6253 | 5888 | 0.76 | \$0.89 |
| South Sudan | LI | AFR | 1080 | 1484 | 0.76 | \$1.53 |
| Sri Lanka | LMI | SEAR | 4516 | 1635 | 0.98 | \$0.61 |
| St. Lucia | UMI | AMR | 14182 | 10 | 0.99 | \$4.55 |
| St. Vincent and the | UMI | AMR | 11501 | 6 | 0.99 | \$1.54 |
| Grenadines |  |  |  |  |  |  |
| Sudan | LI | EMR | 989 | 7646 | 0.48 | \$1.24 |
| Suriname | UMI | AMR | 7431 | 53 | 0.75 | \$0.19 |
| Eswatini | LMI | AFR | 3936 | 142 | 0.91 | \$0.32 |
| Syrian Arab Republic | LI | EMR | 847 | 2273 | 0.81 | \$0.98 |
| Tajikistan | LMI | EUR | 1341 | 1334 | 0.98 | \$3.18 |
| Tanzania | LMI | AFR | 1186 | 10810 | 0.87 | \$3.01 |
| Thailand | UMI | SEAR | 7345 | 3102 | 0.96 | \$5.62 |
| Timor-Leste | LMI | SEAR | 1343 | 157 | 0.89 | \$0.73 |
| Togo | LI | AFR | 1043 | 1342 | 0.95 | \$2.40 |
| Tonga | UMI | WPR | 4864 | 12 | 0.99 | \$1.02 |
| Tunisia | LMI | EMR | 4350 | 902 | 0.97 | \$3.37 |
| Turkiye | UMI | EUR | 15473 | 5746 | 0.99 | \$11.26 |
| Turkmenistan | UMI | EUR | 8572 | 804 | 0.99 | \$4.68 |
| Tuvalu | UMI | WPR | 6345 | 1 | 0.99 | \$0.60 |
| Uganda | LI | AFR | 1073 | 7938 | 0.95 | \$4.88 |
| Ukraine | UMI | EUR | 5389 | 1304 | 0.95 | \$3.49 |
| Uzbekistan | LMI | EUR | 3162 | 4311 | 0.9 | \$3.19 |
| Vanuatu | LMI | WPR | 3543 | 44 | 0.88 | \$0.58 |
| Venezuela | UMI | AMR | 15944 | 2124 | 0.67 | \$0.80 |
| Viet Nam | LMI | WPR | 4717 | 7178 | 0.99 | \$8.63 |
| Yemen | LI | EMR | 433 | 6294 | 0.53 | \$0.21 |
| Zambia | LMI | AFR | 1235 | 3152 | 0.94 | \$3.18 |
| Zimbabwe | LMI | AFR | 2656 | 2307 | 0.93 | \$3.05 |

**Figure 3.**
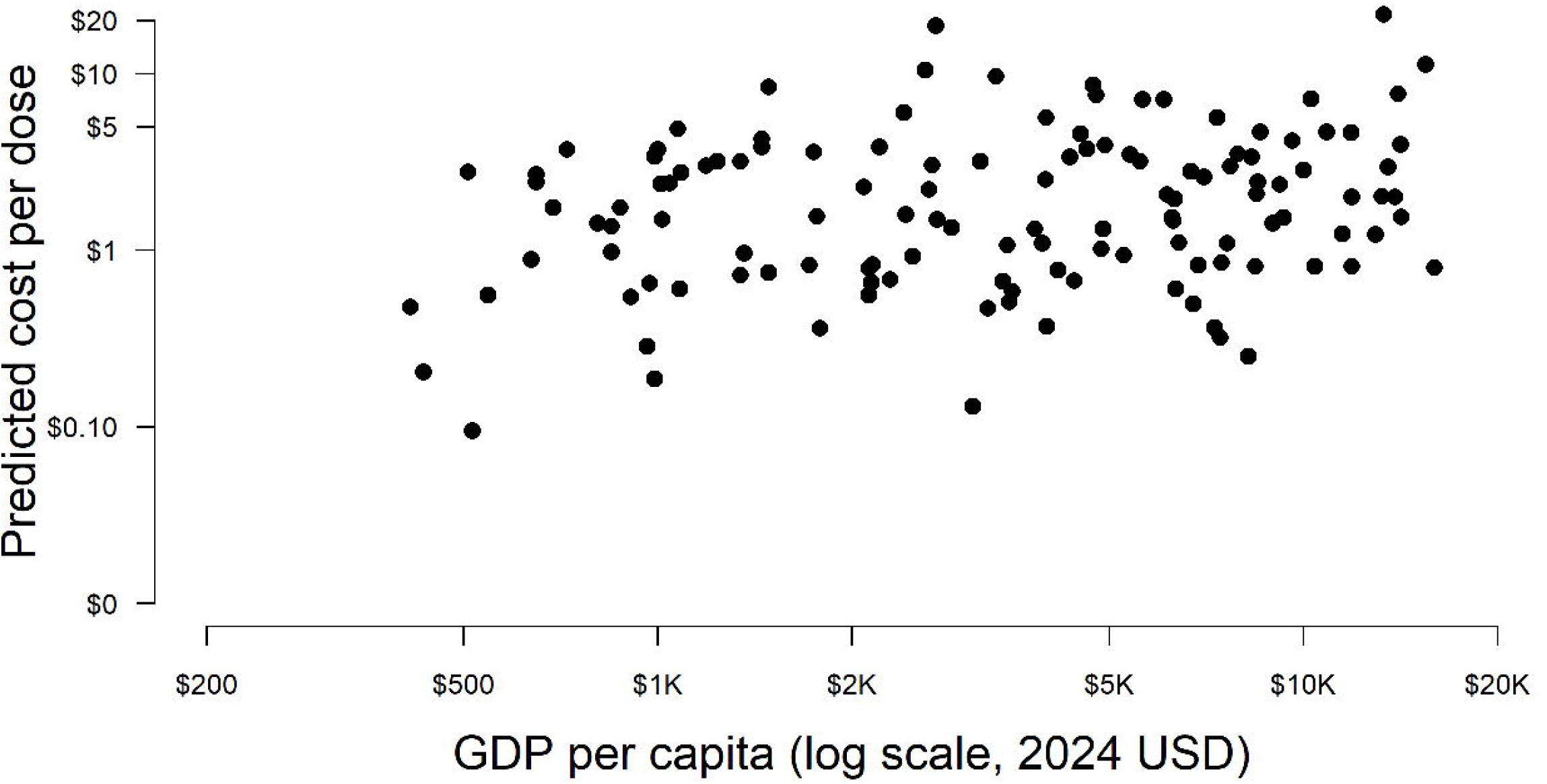
Predicted economic cost per dose in 2024 for routine outreach vaccine delivery by gross domestic product (GDP) per capita for 129 low and middle-income countries.

### Sensitivity analyses

After excluding statistical outliers, the model fit showed certain meaningful changes in coefficient estimates, suggesting that a small number of extreme high-cost observations substantially influenced the original regression estimates (Appendix Table B). The Gamma dispersion parameter alpha increased from 1.412 in the full dataset model to 2.272 after excluding outliers, indicating improved model fit and reduced influence of extreme observations on the residual distribution. This sensitivity analysis resulted in an estimated population-weighted average economic cost per dose of $6.12 ($2.23–13.92) for the full costs of routine outreach vaccine delivery across 129 LMICs, which overlaps with the associated estimates from the primary analysis including outliers, $8.65 ($2.33–23.71). Country-specific economic cost estimates for outreach delivery cost per dose based on the model excluding statistical outliers can be found in Appendix Table C.

## Discussion

For the year 2024, the average economic cost per dose across all LMICs was estimated to be $8.65 ($2.33–23.71) for the full costs of routine outreach vaccine delivery, excluding vaccine costs. These estimates are consistent with the empirical estimates reported in a recent scoping review for the costs of interventions to reach zero-dose children, which ranged from $0.04 to $67 per dose (Levin et al., 2024, UNICEF, 2024), but are notably greater than estimates of $2.34 ($0.80–5.47) in 2024 USD to deliver routine childhood vaccinations on average (Portnoy et al., 2020). Importantly, the estimates presented here should be interpreted as an approximation of the additional delivery resources potentially required to reach underserved populations through outreach and mobile strategies, rather than as direct estimates of the costs of reaching zero-dose children. These estimates can be useful in providing a broad indication of delivery costs to inform resource allocation to reach currently un- and under-reached populations with needed vaccinations.

Using these predicted routine outreach delivery costs per dose as a proxy for reaching zero-dose children, the costs of fully vaccinating a zero-dose child would be scaled by the number of vaccine doses in the vaccine program. For example, if we assume the vaccine program requires 13 doses (e.g., 3 pentavalent vaccine, 3 oral polio vaccine, 3 pneumococcal conjugate vaccine, 3 rotavirus, 1 measles-containing vaccine) for a child to be considered fully vaccinated, and we assume that zero-dose children can be reached at the average unit cost per dose of outreach delivery, the predicted average economic cost per dose would be multiplied by 13, which would be equivalent to $112.45 ($30.29–308.23) to fully vaccinate a zero-dose child across the study countries. However, this calculation should be interpreted as a simplified heuristic intended to illustrate the potential magnitude of delivery costs rather than as a realistic child-level costing model. In practice, the resources required to fully vaccinate a zero-dose child would depend on the child’s age, opportunities for co-administration of vaccines, visit schedules, and dropout between visits.

An alternative illustrative approach would take into account the observed positive association between coverage and unit costs, and therefore focus on the marginal cost per dose. Under this approach, for a country at 90% DTP1 coverage, the estimated marginal cost per dose of outreach-based delivery is $12.11; multiplied by 13, this would imply a cost of $157.43 to fully vaccinate an unvaccinated child under the same simplifying assumptions.

In the best fit model, the continuous explanatory variables included were GDP per capita, under-five population size, and DTP1 coverage. In our analysis, unit costs were predicted to increase with higher GDP per capita, larger under-five populations, and higher DTP1 coverage. The positive association with GDP per capita may reflect higher input and labor costs in higher-income settings, while the positive associations with under-five population and DTP1 coverage may reflect greater resource requirements associated with larger target populations and more extensive outreach efforts in stronger immunization systems. Finally, we assigned the year of data collection as one of the study-level covariates, which serves as a proxy for time-varying characteristics not captured by other model covariates. Costs were predicted to decrease over time, but the magnitude was small. However, it is important to note that regression coefficients from this model do not have a causal interpretation. Our sensitivity analysis results suggest that the primary substantive conclusions of the model were generally preserved, but that several covariate relationships were sensitive to a small number of influential high-cost observations.

There are several limitations to this analysis. First, the studies we included in the analysis did not have a standardized design, and the resulting heterogeneity was not fully captured by our meta-regression analysis. Therefore, the residual variance could reflect both sampling uncertainty as well as non-sampling error due to methodological heterogeneity. However, given the limited number of available studies, restricting the analysis to only narrowly comparable study designs would have substantially reduced the geographic scope and empirical basis of the analysis. We therefore combined evidence across heterogeneous study types to provide a broader evidence base across LMICs, while attempting to account for key methodological differences through study-level covariates in the meta-regression. The 19 countries included in the dataset might not be representative of the 129 LMICs to which we extrapolated predicted costs. Therefore, estimates will be less reliable for countries with covariate values outside the range observed in the analyzed sample—for example, countries with substantially higher or lower GDP per capita, under-five population size, or DTP1 coverage than those represented in the underlying studies. In the sample used to fit the model, the average GDP per capita was $1743.57 (std $1398.95), the average under-five population size was 16,700 (std 32,900), and the average DTP1 coverage was 92.7% (std 6.8%). Predictions for countries with covariate values broadly within these observed ranges are therefore more reflective of interpolation within the empirical data used for model fitting, whereas predictions for countries with values substantially outside these ranges rely more heavily on extrapolation beyond the observed data and should be interpreted with greater caution. In particular, only two observations from currently-defined upper middle-income countries were included in the dataset used for model fitting. Second, this analysis assumed that the estimated unit costs of vaccine delivery are independent of the number of doses in the immunization program. However, we would expect different costs for vaccines requiring unique immunization visits compared to vaccines that can be administered in the same immunization visit. Finally and importantly, outreach and mobile efforts to reach children already reached by ongoing immunization efforts might not be representative of efforts required to fully vaccinate zero-dose children, who are not reached by the programs informing this analysis by definition. While outreach and mobile delivery strategies are likely to play an important role in efforts to reduce the number of zero-dose children, the costs of outreach delivery should not be considered equivalent to the full costs of identifying and reaching zero-dose populations. We expect that fully vaccinating zero-dose children may be even more costly, not only because they are among the most difficult populations to reach, but also because identifying and locating zero-dose children can itself require substantial additional resources. Zero-dose children are often geographically dispersed, mobile, marginalized, or outside formal health system contact, making them challenging to identify through routine service delivery mechanisms. Future studies that focus on the costs of reaching zero dose children should include the costs of identifying and locating those children to ensure that cost estimates reflect the full resource burden required. In particular, we expect higher costs in geographically hard-to-reach or conflict-affected settings, which likewise increase the risk of a child being zero-dose. Additional activities beyond vaccine delivery itself—including community engagement, population mapping, surveillance, and identification of previously unreached children—may contribute importantly to the total resources required to reduce zero-dose prevalence. Our cross-sectional analysis suggests that costs increase as coverage increases; if this relationship holds true longitudinally as individual countries increase coverage, then the average cost per dose would increase significantly as countries reach more children.

In light of these limitations, this analysis may not fully address the decision- and policy-making needs to improve immunization coverage and equity, and reach the Immunization Agenda 2030 goals. However, the need to make policy choices based on imperfect information is unavoidable, and the estimates we report provide an additional evidence source for budgeting and planning around closing immunization coverage gaps.

Our study provides estimates produced via meta-regression analyses that can help countries to improve budgeting and planning for implementation of future interventions to reach zero-dose children, offering critical insights for global health stakeholders and policymakers. These estimates are best interpreted as approximate indicators of the costs associated with outreach- and mobile-based delivery approaches that may contribute to improving coverage among underserved populations, including zero-dose children. Bridging the immunization gap for zero-dose children through routine systems not only advances universal health coverage but also strengthens health systems and improves population resilience to future public health threats (Oyugi et al., 2025). As countries and partners mobilize resources to close this immunization gap, these cost estimates serve as a valuable foundation for strategic planning, prioritization, and sustainable financing.

## Data Availability

All data produced in the present study are available upon reasonable request to the authors

## References

Brenzel, L., Young, D. & Walker, D. G. 2015. Costs and financing of routine immunization: Approach and selected findings of a multi-country study (EPIC). Vaccine, 33 Suppl 1, A13–20.

Clarke-Deelder, E., Suharlim, C., Chatterjee, S., Portnoy, A., Brenzel, L., Ray, A., Cohen, J. L., Menzies, N. A. & Resch, S. C. 2024. Health impact and cost-effectiveness of expanding routine immunization coverage in India through Intensified Mission Indradhanush. Health Policy Plan, 39, 583–592.

Gelman, A. 2006. Prior distributions for variance parameters in hierarchical models. Bayesian Analysis, 1, 515–534.

Gelman, A. 2025. Prior Choice Recommendations. Last updated: 27 April 2025. [Online] Accessed 25 May 2025. Available at: https://github.com/stan-dev/stan/wiki/Prior-Choice-Recommendations.

Hoffman, M. D. & Gelman, A. 2014. The No-U-Turn Sampler: Adaptively Setting Path Lengths in Hamiltonian Monte Carlo. Journal Of Machine Learning Research, 15, 1593–1623.

Hogan, D. & Gupta, A. 2023. Why Reaching Zero-Dose Children Holds the Key to Achieving the Sustainable Development Goals. Vaccines (Basel), 11.

Immunization Costing Action Network (Ican) 2024. Immunization delivery cost catalogue (IDCC). ThinkWell. Last updated: 20 Jun 2024. [Online] Accessed 20 Jun 2024. Available at: https://immunizationeconomics.org/thinkwell-idcc/

Levin, A., Fisseha, T., Reynolds, H. W., CorrÊA, G., Mengistu, T. & Vollmer,N. 2024. Scoping Review of Current Costing Literature on Interventions to Reach Zero-Dose Children in Low-and Middle-Income Countries. Vaccines (Basel), 12.

Oyugi, B., Kallander, K. & Shahabuddin, A. S. M. 2025. Strengthening Primary Health Care Through Implementation Research: Strategies for Reaching Zero-Dose Children in Low-and Middle-Income Countries’ Immunization Programs. Vaccines (Basel), 13.

Portnoy, A., Resch, S. C., Suharlim, C., Brenzel, L. & Menzies, N. A. 2021. What We Do Not Know About the Costs of Immunization Programs in Low-and Middle-Income Countries. Value Health, 24, 67–69.

Portnoy, A., Vaughan, K., Clarke-Deelder, E., Suharlim, C., Resch, S. C., Brenzel, L. & Menzies, N. A. 2020. Producing Standardized Country-Level Immunization Delivery Unit Cost Estimates. Pharmacoeconomics, 38, 995–1005.

Unicef 2024. Costs of Fully Vaccinating a Child. August 2024. [Online] Accessed 25 September 2025. Available at: https://www.unicef.org/documents/costs-fully-vaccinating-child.

United Nations 2024. World Population Prospects. United Nations population division. Last updated: 15 Jul 2024. [Online] Accessed 7 Nov 2024. Available at: https://population.un.org/wpp/

Vaughan, K., Ozaltin, A., Mallow, M., Moi, F., Wilkason, C., Stone, J. & Brenzel, L. 2019. The costs of delivering vaccines in low-and middle-income countries: Findings from a systematic review. Vaccine X, 2, 100034.

Wendt, A., Santos, T. M., Cata-Preta, B. O., Arroyave, L., Hogan, D. R., Mengistu, T., Barros, A. J. D. & Victora, C. G. 2022. Exposure of Zero-Dose Children to Multiple Deprivation: Analyses of Data from 80 Low-and Middle-Income Countries. Vaccines (Basel), 10.

World Bank 2025a. World Bank Country and Lending Groups. Washington, DC: The World Bank. Last updated: 1 July 2025. [Online] Accessed 18 Sep 2025. Available at: https://datahelpdesk.worldbank.org/knowledgebase/articles/906519.

World Bank 2025b. World development indicators. Last updated: 1 Jul 2025. [Online] Accessed 18 Sep 2025. Available at: https://data.worldbank.org/

World Health Organization 2020. Immunization Agenda 2030: a global strategy to leave no one behind. Last updated: 1 April 2020. [Online] Accessed 6 June 2025. Available at: https://www.who.int/publications/m/item/immunization-agenda-2030-a-global-strategy-to-leave-no-one-behind.

World Health Organization 2023. The Big Catch-Up: An Essential Immunization Recovery Plan for 2023 and Beyond. Last Updated; 26 July 2023. [Online] Accessed 6 June 2025. Available at: https://www.who.int/publications/i/item/9789240075511.

World Health Organization 2025. WHO-UNICEF estimates of DTP1 coverage: monitoring system 2024 global summary. Last updated: 15 Jul 2025. [Online] Accessed 18 Sep 2025. Available at: https://immunizationdata.who.int/global

