## Supplementary Appendix for "Approximating vaccine delivery costs to reach zero-dose children: a Bayesian meta-regression analysis"

**Appendix A. Characteristics of the included studies.**

| **Country** | **Year** | **Economic or financial cost** | **Full or incremental costing study** | **Routine or SIA delivery modality** | **Vaccine(s) includes** | **Outreach delivery cost per dose**  **(2024 USD)** | **Source** |
| --- | --- | --- | --- | --- | --- | --- | --- |
| Nigeria | 2016 | Economic | Full | SIA | Measles | 0.09 | [1] |
| Indonesia | 2000 | Economic | Incremental | Routine | HepB | 0.13 | [2] |
| Cameroon | 2011 | Financial | Full | SIA | Measles | 0.16 | [3] |
| India | 2017 | Financial | Incremental | SIA | TCV | 0.43 | [4] |
| Malawi | 2019 | Financial | Incremental | Routine | TCV | 0.52 | [5] |
| Malawi | 2022 | Financial | Incremental | Routine | Malaria | 0.53 | [6] |
| Sierra Leone | 2019 | Financial | Full | SIA | MR OPV | 0.53 | [7] |
| Burkina Faso | 2021 | Financial | Incremental | Routine | Malaria | 0.65 | [8] |
| Benin | 2011 | Economic | Full | Routine | Measles | 0.71 | [9] |
| Kenya | 2022 | Financial | Incremental | Routine | Malaria | 0.72 | [6] |
| Malawi | 2022 | Economic | Incremental | Routine | Malaria | 0.74 | [6] |
| Indonesia | 2016 | Financial | Full | Routine | BCG Measles HepB DTP-HepB-Hib OPV | 0.75 | [10] |
| Burkina Faso | 2021 | Economic | Incremental | Routine | Malaria | 0.78 | [8] |
| Benin | 2011 | Economic | Full | SIA | Measles | 0.82 | [9] |
| Malawi | 2019 | Financial | Incremental | Routine | TCV | 0.88 | [5] |
| Ghana | 2022 | Financial | Incremental | Routine | Malaria | 0.91 | [6] |
| Indonesia | 2016 | Financial | Full | Routine | BCG Measles HepB DTP-HepB-Hib OPV | 0.93 | [10] |
| Malawi | 2019 | Financial | Incremental | SIA | TCV | 0.94 | [5] |
| Sierra Leone | 2019 | Economic | Full | SIA | MR OPV | 1.13 | [7] |
| Kenya | 2022 | Economic | Incremental | Routine | Malaria | 1.26 | [6] |
| Mali | 2021 | Financial | Incremental | Routine | Malaria | 1.32 | [8] |
| Indonesia | 2016 | Financial | Full | Routine | BCG Measles HepB DTP-HepB-Hib OPV | 1.35 | [10] |
| Mali | 2021 | Economic | Incremental | Routine | Malaria | 1.42 | [8] |
| Malawi | 2019 | Economic | Incremental | Routine | TCV | 1.44 | [5] |
| Ghana | 2022 | Economic | Incremental | Routine | Malaria | 1.69 | [6] |
| India | 2017 | Economic | Incremental | SIA | TCV | 1.70 | [4] |
| Malawi | 2019 | Economic | Incremental | Routine | TCV | 1.82 | [5] |
| Uganda | 2015 | Financial | Full | Routine | BCG Measles TT DTP-HepB-Hib OPV IPV PCV10 HPV | 1.83 | [11] |
| Zambia | 2011 | Economic | Full | Routine | BCG Measles DTP-HepB-Hib OPV | 1.86 | [12, 13] |
| Malawi | 2019 | Economic | Incremental | SIA | TCV | 1.88 | [5] |
| Cote d'Ivoire | 2006 | Economic | Full | Routine | BCG DTP-Hib OPV | 11.07 | [14] |
| Cote d'Ivoire | 2006 | Economic | Full | Routine | BCG DTP-Hib OPV | 11.56 | [14] |
| Malawi | 2019 | Financial | Incremental | Routine | Malaria | 2.15 | [15] |
| Tanzania | 2016 | Financial | Full | Routine | BCG MR DTP-HepB-Hib OPV Rotavirus (2 doses) PCV13 | 2.23 | [16] |
| Kenya | 2019 | Financial | Incremental | Routine | Malaria | 2.6 | [15] |
| Cameroon | 2009 | Economic | Full | Routine | BCG Measles DTP-HepB-Hib OPV YF | 2.68 | [17] |
| Ghana | 2019 | Financial | Incremental | Routine | Malaria | 2.92 | [15] |
| Malawi | 2019 | Economic | Incremental | Routine | Malaria | 3.15 | [15] |
| Indonesia | 2016 | Financial | Full | Routine | BCG Measles HepB DTP-HepB-Hib OPV | 3.72 | [10] |
| Viet Nam | 2017 | Financial | Full | SIA | Td | 4.43 | [18] |
| Tanzania | 2016 | Economic | Full | Routine | BCG MR DTP-HepB-Hib OPV Rotavirus (2 doses) PCV13 | 4.71 | [19] |
| Mexico | 2002 | Economic | Full | SIA | OPV | 4.72 | [20] |
| Kenya | 2019 | Economic | Incremental | Routine | Malaria | 4.88 | [15] |
| India | 2017 | Economic | Incremental | Routine | BCG Measles MR HepB DTP Td TT DTwP-HepB-Hib OPV IPV Rotavirus (3 doses) PCV13 JE | 5.01 | [21] |
| Ghana | 2019 | Economic | Incremental | Routine | Malaria | 5.18 | [15] |
| Iraq | 2018 | Economic | Full | Routine | BCG Measles MMR HepB DTwP DTP-HepB-Hib OPV IPV Rotavirus (2 doses) PCV13 | 5.60 | [22] |
| India | 2017 | Economic | Incremental | Routine | DTP | 66.26 | [21] |
| Honduras | 2011 | Economic | Full | Routine | BCG MMR HepB DTP-HepB-Hib OPV Rotavirus (2 doses) PCV13 YF | 9.80 | [13, 23] |

**Appendix Table B. Results for regressions of outreach delivery unit cost per dose on predictors, excluding statistical outliers.**

| Variable | Mean coefficient (standard error) |
| --- | --- |
| Intercept | - 0.218 (0.004) |
| Year | - 0.131 (0.002) |
| Economic cost indicator [$Econ$] | 0.546 (0.002) |
| Full costing indicator [$Full$] | 0.261 (0.003) |
| Routine delivery indicator [$Routine$] | 0.458 (0.003) |
| log(GDP) | 0.288 (0.001) |
| log(Pop) | 0.056 (0.001) |
| DTP1 coverage | 0.460 (0.001) |
| Gamma dispersion parameter alpha | 2.272 |

**Appendix C. Low- and middle-income country parameters with predicted economic outreach delivery cost per dose in 2024, based on model fit excluding statistical outliers.**

| **Country** | **World Bank Income Level^a^ [9]** | **WHO Region** | **Per-Capita Gross Domestic Product [13]** | **Under-Five Population (thousands) [13]** | **DTP1 Coverage [14]** | **Predicted Economic Outreach Delivery Cost per Dose for Childhood Vaccines** |
| --- | --- | --- | --- | --- | --- | --- |
| Afghanistan | LI | EMR | 414 | 6656 | 0.66 | $0.45 |
| Albania | UMI | EUR | 10012 | 145 | 0.99 | $13.68 |
| Algeria | UMI | AFR | 5631 | 4753 | 0.98 | $9.96 |
| Angola | LMI | AFR | 2122 | 6219 | 0.7 | $1.10 |
| Argentina | UMI | AMR | 13858 | 2751 | 0.8 | $4.82 |
| Armenia | UMI | EUR | 8501 | 177 | 0.97 | $10.96 |
| Azerbaijan | UMI | EUR | 7284 | 675 | 0.65 | $1.40 |
| Bangladesh | LMI | SEAR | 2593 | 16505 | 0.99 | $7.98 |
| Belarus | UMI | EUR | 8317 | 399 | 0.98 | $11.54 |
| Belize | UMI | AMR | 8430 | 36 | 0.89 | $6.69 |
| Benin | LMI | AFR | 1485 | 2176 | 0.76 | $1.30 |
| Bhutan | LMI | SEAR | 3839 | 48 | 0.98 | $7.90 |
| Bolivia | LMI | AMR | 4001 | 1251 | 0.65 | $1.05 |
| Bosnia and Herzegovina | UMI | EUR | 8957 | 130 | 0.91 | $7.65 |
| Botswana | UMI | AFR | 7695 | 294 | 0.98 | $11.09 |
| Brazil | UMI | AMR | 10280 | 13361 | 0.91 | $8.71 |
| Burkina Faso | LI | AFR | 987 | 3351 | 0.95 | $3.73 |
| Burundi | LI | AFR | 154 | 2167 | 0.89 | $1.22 |
| Cabo Verde | UMI | AFR | 5273 | 39 | 0.93 | $6.72 |
| Cambodia | LMI | WPR | 2628 | 1816 | 0.9 | $4.01 |
| Cameroon | LMI | AFR | 1762 | 4393 | 0.82 | $2.08 |
| Central African Republic | LI | AFR | 516 | 941 | 0.52 | $0.20 |
| Chad | LI | AFR | 1016 | 3547 | 0.84 | $1.86 |
| China | UMI | WPR | 13303 | 58042 | 0.97 | $15.71 |
| Colombia | UMI | AMR | 7914 | 3553 | 0.89 | $6.45 |
| Comoros | LMI | AFR | 1784 | 114 | 0.8 | $1.70 |
| Congo | LMI | AFR | 2482 | 886 | 0.82 | $2.30 |
| Congo DR | LI | AFR | 647 | 19276 | 0.82 | $1.52 |
| Cote d'Ivoire | LMI | AFR | 2710 | 4594 | 0.8 | $2.23 |
| Cuba | UMI | AMR | 9605 | 504 | 0.99 | $13.30 |
| Djibouti | LMI | EMR | 3496 | 115 | 0.82 | $2.67 |
| Dominica | UMI | AMR | 10405 | 4 | 0.96 | $13.00 |
| Dominican Republic | UMI | AMR | 10876 | 1004 | 0.97 | $12.46 |
| Ecuador | UMI | AMR | 6875 | 1404 | 0.74 | $2.32 |
| Egypt | LMI | EMR | 3338 | 11781 | 0.99 | $8.73 |
| El Salvador | UMI | AMR | 5580 | 496 | 0.98 | $9.44 |
| Equatorial Guinea | UMI | AFR | 6745 | 252 | 0.75 | $2.42 |
| Eritrea | LI | AFR | 689 | 461 | 0.97 | $3.40 |
| Ethiopia | LI | AFR | 1011 | 19067 | 0.81 | $5.01 |
| Fiji | UMI | WPR | 6288 | 83 | 0.95 | $1.69 |
| Gabon | UMI | AFR | 8219 | 331 | 0.62 | $8.28 |
| Gambia | LI | AFR | 909 | 384 | 0.82 | $1.26 |
| Georgia | UMI | EUR | 9194 | 242 | 0.95 | $1.44 |
| Ghana | LMI | AFR | 2406 | 4226 | 0.99 | $9.99 |
| Grenada | UMI | AMR | 11872 | 7 | 0.93 | $7.20 |
| Guatemala | UMI | AMR | 6150 | 1879 | 0.86 | $11.16 |
| Guinea | LMI | AFR | 1717 | 2195 | 0.77 | $4.63 |
| Guinea-Bissau | LI | AFR | 963 | 297 | 0.74 | $1.47 |
| Haiti | LMI | AMR | 2143 | 1236 | 0.76 | $0.90 |
| Honduras | LMI | AMR | 3426 | 1138 | 0.75 | $1.50 |
| India | LMI | SEAR | 2697 | 114665 | 0.96 | $1.75 |
| Indonesia | UMI | SEAR | 4925 | 22349 | 0.83 | $7.62 |
| Iran | UMI | EMR | 4771 | 6201 | 0.98 | $3.79 |
| Iraq | UMI | EMR | 6074 | 5560 | 0.97 | $9.31 |
| Jamaica | UMI | AMR | 7020 | 166 | 0.99 | $9.71 |
| Jordan | LMI | EMR | 4618 | 1163 | 0.97 | $11.34 |
| Kazakhstan | UMI | EUR | 14005 | 2126 | 0.99 | $8.16 |
| Kenya | LMI | AFR | 2206 | 7037 | 0.91 | $16.36 |
| Kiribati | LMI | WPR | 2289 | 16 | 0.96 | $4.18 |
| Korea DPR | LI | SEAR | 1447 | 1711 | 0.99 | $5.44 |
| Kyrgyz Republic | LMI | EUR | 2419 | 788 | 0.9 | $5.56 |
| Lao PDR | LMI | WPR | 2124 | 796 | 0.76 | $3.77 |
| Lebanon | LMI | EMR | 3478 | 452 | 0.86 | $1.48 |
| Lesotho | LMI | AFR | 972 | 266 | 0.86 | $3.44 |
| Liberia | LI | AFR | 846 | 773 | 0.91 | $1.89 |
| Libya | UMI | EMR | 6318 | 644 | 0.9 | $2.53 |
| North Macedonia | UMI | EUR | 9310 | 95 | 0.93 | $5.97 |
| Madagascar | LI | AFR | 545 | 4588 | 0.7 | $0.62 |
| Malawi | LI | AFR | 508 | 3080 | 0.94 | $2.67 |
| Malaysia | UMI | WPR | 11867 | 2271 | 0.93 | $10.13 |
| Maldives | UMI | SEAR | 13216 | 31 | 0.99 | $16.50 |
| Mali | LI | AFR | 1086 | 4158 | 0.91 | $3.02 |
| Marshall Islands | UMI | WPR | 7467 | 4 | 0.98 | $12.18 |
| Mauritania | LMI | AFR | 2083 | 800 | 0.95 | $4.89 |
| Mauritius | UMI | AFR | 11872 | 61 | 0.97 | $13.35 |
| Mexico | UMI | AMR | 14158 | 10309 | 0.83 | $6.07 |
| Micronesia | LMI | WPR | 4166 | 12 | 0.96 | $7.46 |
| Moldova | UMI | EUR | 7618 | 179 | 0.87 | $5.42 |
| Mongolia | UMI | WPR | 6691 | 353 | 0.97 | $9.67 |
| Montenegro | UMI | EUR | 12935 | 37 | 0.92 | $10.26 |
| Morocco | LMI | EMR | 3993 | 3180 | 0.98 | $8.37 |
| Mozambique | LI | AFR | 647 | 5669 | 0.9 | $2.34 |
| Myanmar | LMI | SEAR | 1359 | 4432 | 0.76 | $1.29 |
| Namibia | LMI | AFR | 4413 | 409 | 0.79 | $2.49 |
| Nepal | LMI | SEAR | 1447 | 2792 | 0.98 | $5.31 |
| Nicaragua | LMI | AMR | 2848 | 657 | 0.88 | $3.57 |
| Niger | LI | AFR | 723 | 4667 | 0.95 | $3.35 |
| Nigeria | LMI | AFR | 807 | 33527 | 0.71 | $0.87 |
| Pakistan | LMI | EMR | 1485 | 31789 | 0.94 | $8.93 |
| Papua New Guinea | LMI | WPR | 3076 | 1229 | 0.49 | $4.74 |
| Paraguay | UMI | AMR | 6416 | 679 | 0.82 | $0.38 |
| Peru | UMI | AMR | 8452 | 2692 | 0.83 | $3.63 |
| Philippines | LMI | WPR | 3985 | 9602 | 0.82 | $4.52 |
| Rwanda | LI | AFR | 1000 | 1878 | 0.99 | $3.10 |
| Samoa | UMI | WPR | 4899 | 28 | 0.99 | $4.77 |
| Sao Tome and Principe | LMI | AFR | 3245 | 31 | 0.87 | $9.66 |
| Senegal | LMI | AFR | 1744 | 2452 | 0.96 | $3.58 |
| Serbia | UMI | EUR | 13524 | 315 | 0.95 | $5.00 |
| Sierra Leone | LI | AFR | 873 | 1164 | 0.92 | $12.24 |
| Solomon Islands | LMI | WPR | 2149 | 104 | 0.91 | $2.78 |
| Somalia | LI | EMR | 637 | 3412 | 0.78 | $3.73 |
| South Africa | UMI | AFR | 6253 | 5888 | 0.76 | $1.05 |
| South Sudan | LI | AFR | 1080 | 1484 | 0.76 | $2.59 |
| Sri Lanka | LMI | SEAR | 4516 | 1635 | 0.98 | $1.11 |
| St. Lucia | UMI | AMR | 14182 | 10 | 0.99 | $8.69 |
| St. Vincent and the Grenadines | UMI | AMR | 11501 | 6 | 0.99 | $17.95 |
| Sudan | LI | EMR | 989 | 7646 | 0.48 | $16.29 |
| Suriname | UMI | AMR | 7431 | 53 | 0.75 | $0.23 |
| Eswatini | LMI | AFR | 3936 | 142 | 0.91 | $2.61 |
| Syrian Arab Republic | LI | EMR | 847 | 2273 | 0.81 | $1.40 |
| Tajikistan | LMI | EUR | 1341 | 1334 | 0.98 | $4.99 |
| Tanzania | LMI | AFR | 1186 | 10810 | 0.87 | $2.55 |
| Thailand | UMI | SEAR | 7345 | 3102 | 0.96 | $9.78 |
| Timor-Leste | LMI | SEAR | 1343 | 157 | 0.89 | $2.63 |
| Togo | LI | AFR | 1043 | 1342 | 0.95 | $3.67 |
| Tonga | UMI | WPR | 4864 | 12 | 0.99 | $9.87 |
| Tunisia | LMI | EMR | 4350 | 902 | 0.97 | $7.89 |
| Turkiye | UMI | EUR | 15473 | 5746 | 0.99 | $17.58 |
| Turkmenistan | UMI | EUR | 8572 | 804 | 0.99 | $12.58 |
| Tuvalu | UMI | WPR | 6345 | 1 | 0.99 | $12.82 |
| Uganda | LI | AFR | 1073 | 7938 | 0.95 | $4.05 |
| Ukraine | UMI | EUR | 5389 | 1304 | 0.95 | $7.72 |
| Uzbekistan | LMI | EUR | 3162 | 4311 | 0.9 | $4.49 |
| Vanuatu | LMI | WPR | 3543 | 44 | 0.88 | $3.97 |
| Venezuela | UMI | AMR | 15944 | 2124 | 0.67 | $2.38 |
| Viet Nam | LMI | WPR | 4717 | 7178 | 0.99 | $9.97 |
| Yemen | LI | EMR | 433 | 6294 | 0.53 | $0.22 |
| Zambia | LMI | AFR | 1235 | 3152 | 0.94 | $3.82 |
| Zimbabwe | LMI | AFR | 2656 | 2307 | 0.93 | $4.93 |

^a^ LI: Gross national income (GNI) per capita of $1,135 or less in 2024; LMI: GNI per capita $1,136 and $4,4915; UMI: GNI per capita of $4,496 and $13,935.

Note: AFR = African region; AMR = Region of the Americas; DTP1 = diphtheria-tetanus-pertussis first dose coverage; EMR = Eastern Mediterranean region; EUR = European region; LI = low-income; LMI = lower middle-income; SEAR = Southeast Asian region; UMI = upper middle-income; WPR = Western Pacific region.

**References**

1.	Sibeudu FT, Onwujekwe OE, Okoronkwo IL. Cost analysis of supplemental immunization activities to deliver measles immunization to children in Anambra state, south-east Nigeria. Vaccine. 2020;38(37):5947-54. Epub 20200707. doi: 10.1016/j.vaccine.2020.06.072. PubMed PMID: 32651114.

2.	Levin CE, Nelson CM, Widjaya A, Moniaga V, Anwar C. The costs of home delivery of a birth dose of hepatitis B vaccine in a prefilled syringe in Indonesia. Bull World Health Organ. 2005;83(6):456-61. Epub 20050617. PubMed PMID: 15976897; PubMed Central PMCID: PMCPMC2626261.

3.	Sume GE, Fouda AA, Kobela M, Nguelé S, Emah I, Atem P. A locally initiated and executed measles outbreak response immunization campaign in the nylon health district, Douala Cameroon 2011. BMC Res Notes. 2013;6:100. Epub 20130316. doi: 10.1186/1756-0500-6-100. PubMed PMID: 23497712; PubMed Central PMCID: PMCPMC3607843.

4.	Song D, Pallas SW, Shimpi R, Ramaswamy N, Haldar P, Harvey P, et al. Delivery cost of the first public sector introduction of typhoid conjugate vaccine in Navi Mumbai, India. PLOS Glob Public Health. 2023;3(1):e0001396. Epub 20230104. doi: 10.1371/journal.pgph.0001396. PubMed PMID: 36962873; PubMed Central PMCID: PMCPMC10022355.

5.	Debellut F, Mkisi R, Masoo V, Chisema M, Mwagomba D, Mtenje M, et al. Projecting the cost of introducing typhoid conjugate vaccine (TCV) in the national immunization program in Malawi using a standardized costing framework. Vaccine. 2022;40(12):1741-6. Epub 20220210. doi: 10.1016/j.vaccine.2022.02.016. PubMed PMID: 35153097; PubMed Central PMCID: PMCPMC8917043.

6.	Baral R, Levin A, Odero C, Pecenka C, Tabu C, Mwendo E, et al. Costs of continuing RTS,S/ASO1E malaria vaccination in the three malaria vaccine pilot implementation countries. PLoS One. 2021;16(1):e0244995. Epub 20210111. doi: 10.1371/journal.pone.0244995. PubMed PMID: 33428635; PubMed Central PMCID: PMCPMC7799756.

7.	Immunization Costing Action Network (ICAN). Estimating the Cost of the integrated Measles-Rubella Campaign in Sierra Leone. Washington, DC: ThinkWell. 2021.

8.	Diawara H, Bocoum FY, Dicko A, Levin A, Lee C, Koita F, et al. Cost of introducing and delivering malaria vaccine (RTS,S/AS01(E)) in areas of seasonal malaria transmission, Mali and Burkina Faso. BMJ Glob Health. 2023;8(4). doi: 10.1136/bmjgh-2022-011316. PubMed PMID: 37068848; PubMed Central PMCID: PMCPMC10111920.

9.	Kaucley L, Levy P. Cost-effectiveness analysis of routine immunization and supplementary immunization activity for measles in a health district of Benin. Cost Eff Resour Alloc. 2015;13:14. Epub 20150820. doi: 10.1186/s12962-015-0039-7. PubMed PMID: 26300696; PubMed Central PMCID: PMCPMC4545866.

10.	Immunization Costing Action Network (ICAN). The Cost of Delivering Vaccines Using Different Delivery Strategies in High Coverage Areas in Indonesia. Washington, DC: ThinkWell. 2019.

11.	Chebrot I, Kisakye A, Kwesiga B, Okello D, Kiiza D, Kabwongera E, et al. The Cost of Routine Immunization Services in A Poor Urban Setting in Kampala, Uganda: Findings of a Facility-Based Costing Study. Journal of Immunological Sciences ,. 2018;2(SI1):94-102.

12.	Schütte C, Chansa C, Marinda E, Guthrie TA, Banda S, Nombewu Z, et al. Cost analysis of routine immunisation in Zambia. Vaccine. 2015;33 Suppl 1:A47-52. doi: 10.1016/j.vaccine.2014.12.040. PubMed PMID: 25919174.

13.	Suharlim C, Menzies NA. Personal communication. 2018.

14.	Douba A, Dagnan SN, Zengbe-Acray P, Aka J, Lépri-Aka N. [Estimated costs of the expanded program of immunization in the health district of Grand Bassam, Cote d'Ivoire]. Sante Publique. 2011;23(2):113-21. PubMed PMID: 21896225.

15.	Baral R, Levin A, Odero C, Pecenka C, Tanko Bawa J, Antwi-Agyei KO, et al. Cost of introducing and delivering RTS,S/AS01 malaria vaccine within the malaria vaccine implementation program. Vaccine. 2023;41(8):1496-502. Epub 20230127. doi: 10.1016/j.vaccine.2023.01.043. PubMed PMID: 36710234; PubMed Central PMCID: PMCPMC9946791.

16.	Immunization Costing Action Network (ICAN). The Costs of Different Vaccine Delivery Strategies to Reach Children Up to 18 Months in Rural and Urban Areas in Tanzania. Washington, DC: ThinkWell. 2019.

17.	Ebong CE, Levy P. Impact of the introduction of new vaccines and vaccine wastage rate on the cost-effectiveness of routine EPI: lessons from a descriptive study in a Cameroonian health district. Cost Eff Resour Alloc. 2011;9(1):9. Epub 20110528. doi: 10.1186/1478-7547-9-9. PubMed PMID: 21619674; PubMed Central PMCID: PMCPMC3123280.

18.	Mai VQ, Boonstoppel L, Vaughan K, Schutte C, Ozaltin A, Hong DT, et al. Cost of Delivering Tetanus Toxoid and Tetanus-Diphtheria Vaccination in Vietnam and the Budget Impact of Proposed Changes to the Schedule. Glob Health Sci Pract. 2022;10(2). Epub 20220429. doi: 10.9745/ghsp-d-21-00482. PubMed PMID: 35487560; PubMed Central PMCID: PMCPMC9053159.

19.	Vaughan K, Clarke-Deelder E, Tani K, Lyimo D, Mphuru A, Manzi F, et al. Immunization costs, from evidence to policy: Findings from a nationally representative costing study and policy translation effort in Tanzania. Vaccine. 2020;38(48):7659-67. Epub 20201017. doi: 10.1016/j.vaccine.2020.10.004. PubMed PMID: 33077300; PubMed Central PMCID: PMCPMC7604567.

20.	Mascareñas A, Salinas J, Tasset-Tisseau A, Mascareñas C, Khan MM. Polio immunization policy in Mexico: economic assessment of current practice and future alternatives. Public Health. 2005;119(6):542-9. doi: 10.1016/j.puhe.2004.08.020. PubMed PMID: 15826896.

21.	Clarke-Deelder E, Suharlim C, Chatterjee S, Portnoy A, Brenzel L, Ray A, et al. Health impact and cost-effectiveness of expanding routine immunization coverage in India through Intensified Mission Indradhanush. Health Policy Plan. 2024;39(6):583-92. doi: 10.1093/heapol/czae024. PubMed PMID: 38590052; PubMed Central PMCID: PMCPMC11145919.

22.	Garcia C, Hossain SM, Lami F, Jabbar F, Rahi A, Kadhim KA, et al. Costs of childhood vaccine delivery in Iraq: a cross-sectional study. BMJ Open. 2022;12(9):e059566. Epub 20220913. doi: 10.1136/bmjopen-2021-059566. PubMed PMID: 36100299; PubMed Central PMCID: PMCPMC9472113.

23.	Pan American Health Organization (PAHO). Comprehensive costing and financial flows analysis of the national immunization program in Honduras, 2011. 2014.
